# Clearing your mind of work-related stress through *moderate-to-vigorous* and *leisure-time* physical activity: What ‘dose’ it takes?

**DOI:** 10.1101/2020.05.11.20097931

**Authors:** Jean-Philippe Lachance, Marc Corbière, Gabriel Hains-Monfette, Paquito Bernard

## Abstract

**Background:** Work is reported as one of the main sources of psychological stress. Because of its role in the onset of burnout and impact on economic and health systems, work-related stress (WS) has become an issue of much concern. Among modifiable factors capable of reducing WS, two categories of physical activity (PA), namely leisure-time and moderate-to-vigorous physical activity (LTPA and MVPA), show promising evidence. Previous findings suggest that LTPA and MVPA allow adults to experience psychological detachment from job demands and restore their depleted resources at work. However, the optimal independent doses of LTPA and MVPA associated with a lower WS level has not yet been established.

**Methods:** The aim of this study was therefore to address this gap using a cross-sectional, nationally representative sample of 4 200 Canadian workers. MVPA was measured through accelerometry and a self-reported assessment was conducted to collect data on WS and LTPA.

**Results:** Generalized additive models indicated that one hour a day on average spent doing a LTPA of 8.5 METs-hour was associated with the highest benefits on WS (p < 0.001, Adjusted R2 = 0.04) while the optimal average daily dose of MVPA was around 90 minutes (p < 0.001, Adjusted R2 = 0.04). Noteworthy, first signs of WS reduction appear long before optimum is reached (e.g. 30 minutes of MVPA), stressing the relevance of merely doing an LTPA/MVPA regardless of the dose.

**Conclusion:** Findings offer practical recommendations for public health policies on the optimal doses of MVPA/LTPA associated with decreased WS.

**What is already known on this subject?:**

- Work-related stress (WS) is one of the major sources of psychological stress and creates fertile ground for the onset and aggravation of physiological and psychiatric conditions.
- Leisure-time and moderate-to-vigorous physical activity (LTPA and MVPA) are known to exert a stress-buffering effect, yet their respective dose-response relationship with WS remains unknown.

**What this study adds?:**

- This is the first study that has employed a nationally representative sample to identify the optimal amount of LTPA and MVPA associated with lower WS levels.
- One hour a day on average spent doing an LTPA of 8.5 METs-hour is associated with the highest benefits, the optimal average daily dose of MVPA is around 90 minutes, and first signs of reduction in WS appear long before optimum are reached, stressing the relevance of merely practising a LTPA/MVPA regardless of the dose.

## BACKGROUND

Psychological stress has been dubbed “the health epidemic of the 21st century” by the World Health Organization.^1^ Psychological stress is classically defined as “a particular relationship between the person and the environment that is appraised by the person as taxing or exceeding his or her resources and endangering his or her well-being”.^2^ While acute punctual stress is unavoidable and adaptive, repeated exposure to perceived threat that results in a significant release of stress hormones may be damaging.^3^ As pioneer studies on stress have pointed out, no organism can be maintained continuously in a state of alarm without entering a stage of exhaustion.^4^ Chronic stress is indeed prospectively associated with the onset and aggravation of metabolic syndrome,^5^ neurodegenerative diseases^6^ as well as psychiatric conditions such as major depression.^7^

Work-related stress (WS) is one of the major sources of psychological stress.^8,9^ In Canada, over one in four workers describe their lives as highly stressful and work is cited as the main source of stress for 60% of those.^8^ In absolute numbers, this prevalence represented in 2010 three millions of people across the country. When unsuccessfully managed at an individual and organizational levels, WS can become fertile ground for burnout,^10^ a costly occupational-related syndrome characterized by feelings of energy depletion or exhaustion, increased mental distance from one’s job, feelings of negativism or cynicism related to one’s job and reduced professional efficacy.^11^

Over the recent decades, life habits such as physical activity (PA) have been receiving much attention from scientists in search of modifiable protective factors against WS. There is scientific evidence showing PA is negatively associated with WS and other health outcomes related with WS such as depression.^12^ One explanation for that protective effect lies in the idea that PA consists of a recovery activity allowing the person to experience psychological detachment from the job demands and restore their depleted resources at work.^13^ It is also well established that the neurophysiological correlates of PA such as the production of endorphins improve mood.^14^

In line with these hypotheses, a growing body of evidence suggests that leisure-time PA (LTPA) as well as moderate-to-vigorous PA (MVPA) are particularly protective against WS.^15,16^ Indeed, some PA modalities such as the domain (occupational, transportation, housework, leisure time) and the intensity seem to play a moderating role in the relationship between PA and mental health. Two recent meta-analyses^17,18^ have shown that LTPA, contrary to other PA domains such as occupational PA or household PA, has been found to be positively associated with mental health outcomes including WS. Regarding the intensity of PA, studies suggest that daily time spent in MVPA could be effective in improving mental health-related outcomes,^15,16^ and eventually reducing WS.

### Purpose of the Present Study

To date, however, the shape of the association with WS, both for LTPA and MVPA, have not been examined so far in a national representative sample. Without any knowledge of the dose-response relationship (e.g. the dose producing the optimal response), it remains difficult to optimize appropriate LTPA and MVPA interventions to ensure the strongest possible decrease in WS at a population level. By bridging the gap in the literature on PA and WS, the aim of this study was to identify the respective dose-response associations of LTPA and MVPA with WS among a high-quality representative national sample of workers.

## METHODS

### Data sources and dataset description

Data were obtained from the combined cycles 1 and 2 (2007 to 2009 and 2009 to 2011) of the Canadian Health Measures Survey (CHMS),^19^ a nationally representative cross-sectional survey of Canadians led by Statistics Canada in partnership with Health Canada and the Public Health Agency of Canada. Data were collected in two stages. First, sociodemographic and general health information were collected during an interview at the participants’ home. Then, weight and height measurements were collected during a subsequent visit to a mobile clinic. The CHMS cycles 1 and 2 employ a stratified sampling protocol to recruit a representative sample of respondents aged from 6 to 79 years old living in privately occupied dwellings in five provinces (New Brunswick, Quebec, Ontario, Alberta, British Columbia) covering approximately 96% of the Canadian population, east to west, with larger and smaller population densities. People living in one of the three territories (Northwest Territories, Nunavut, and Yukon), Indian Reserves or Crown lands, residents of institutions, full-time members of the Canadian Forces and residents of certain remote regions were excluded for feasibility reasons. This survey was approved by the Health Canada’s Research Ethics Board and all respondents provided written informed consent. Detailed information on background, methodology and ethical issues of the CHMS are available elsewhere.^19,20^

In our study, we included Canadians aged 18 to 65 years who reported being employed in the past 12 months with complete data for self-perceived chronic work-related stress (WS), physical activity (PA) data from accelerometer and self-reported leisure-time physical activity (LTPA). Pregnant women and people with functional limitations were not included in analyses.

### Variables

#### Outcome: Self-perceived work-related stress (WS)

Self-perceived chronic work-related stress was assessed using this five-point Likert scale item: “Would you say that most days at work in the past 12 months were: Not at all stressful (coded as ‘1’), Not very stressful (‘2’), A bit stressful (‘3’), Quite a bit stressful (‘4’), Extremely stressful (‘5’)?” Firstly introduced within the Canadian Community Health Survey, this item is widely used to evaluate the level of self-perceived WS in the Canadian population.^21^

#### Exposure 1: Device-assessed Moderate-to-Vigorous Physical Activity (MVPA)

Upon completion of a mobile examination center visit, participants were asked to wear an Actical accelerometer (Phillips-Respironics, 17 grams, omnidirectional accelerometer) over their right hip on an elasticized belt during their waking hours for 7 consecutive days. The Actical is a valid and reliable instrument to measure PA in adults.^22^ The accelerometers started to collect data the day after the mobile examination centre appointment. The monitors were then posted to Statistics Canada. The data were validated by research assistants to determine if they were still within the manufacturer’s calibration specifications. The Actical measures and records time-stamped acceleration in all directions, thereby indicating the intensity of physical activity. The digitized values are summed over an interval of one minute. Accelerometer data were in a count value per minute (cpm) and also into steps accumulated per minute.^23^ All data are blind to respondents while they are wearing the device. Consistent with common analytical procedures for accelerometry, a valid day was defined as 10 or more hours of wear time and respondents with 4 or more valid days were retained for analyses.^24^ Accelerometer data were not included in the analyses if a participant had extreme counts (i.e. >20 000 cpm).^24^ The number of minutes per day spent in MVPA was categorized using validated cut-offs of cpm for adults, which corresponds to ≥1535 cpm.^23^ The average minutes per day of MVPA was used in analyses.

#### Exposure 2: Daily energy expenditure in Leisure-Time Physical Activities (LTPA)

As a composite variable, LTPA energy expenditure was determined from information about the frequency and duration of respondents’ participation in a variety of physical activities during leisure time by the respondent in the past three months. For each reported activity (a total of 21 leisure activities such as gardening, soccer, swimming, as well as all other non-listed choices of leisure PA - see supplementary material), energy expenditure was calculated by multiplying the number of times a respondent engaged in the activity over a 12-month period (a 3-month recall period multiplied by 4) by the average duration in minutes and the energy cost of the activity (the metabolic equivalent - MET, i.e. kilocalories expended per kilogram of weight per hour of activity). For each LTPA, the MET is a value of metabolic energy cost expressed as a multiple of the resting metabolic rate. For example, an activity of 4 METs requires four times the amount of energy the body needs when it is at rest. To calculate the daily energy expenditure for an activity, the yearly estimate was divided by 365. This calculation was repeated for all leisure-time activities reported, and the resulting estimates were summed to yield average daily energy expenditure. MET values tend to be expressed in three intensity levels (i.e. low, medium, high). Since the CHMS questions did not ask the respondent to specify the intensity level of their activities, the MET values adopted corresponded to the low intensity value of each activity. This approach is adopted from the Canadian Fitness and Lifestyle Research Institute because individuals tend to overestimate the intensity, frequency and duration of their activities (Statistics Canada, 2007). The formula used to derive the energy expenditure in LTPA was:

> Energy expenditure in LTPA = (N * D * MET value) / 365
>
> Where:
>
> N = the number of times a respondent engaged in an activity over a 12-month period
>
> D = the average duration in hours of the activity
>
> MET value = the energy cost of the activity expressed as kilocalories expended per kilogram of body weight per hour of activity (kcal/kg per hour)/365 (to convert yearly data into daily data, namely METs-hour per day)

#### Covariates

Sociodemographic covariates included age, sex (m/w), daily smoking (cotinine blood level in ng/mL), household income (ranged from < 15 000 to 100 000$+), marital status and education levels (ranged from < high school to university). Accelerometer wear time (daily minutes) and season of accelerometer assessment were also computed and included in analyses. Body mass index was computed by measuring an individual’s weight and height during mobile clinic visit. All these covariables are commonly related to PA in adults.^25^ Constructs potentially related to WS such as self-perceived general stress and self-perceived mental health, both measured on a five-point Likert scale, were also included in the models.

### Statistical analysis

Generalized additive models (GAMs) were used to examine the shape of the association between WS and MVPA, and then between WS and LTPA. The GAM is an extension of the generalized linear model in that one or more predictors may be specified using a smooth function.^26^ GAM is a nonparametric model that allows nonlinear relationships to be modelized without specifying the functional form a priori. The GAM is estimated using a penalized maximum likelihood procedure – usually iteratively reweighted least squares. Predictions from GAM were plotted with 95% confidence intervals. To account for the complex, multistage probability sampling design, the weights provided by the CHMS were used in the analyses (i.e. activity monitor subsample weights combining cycles 1 and 2). All analyses were performed using survey packages^27^ in R version 3.3.

## RESULTS

### Sample characteristics

Accelerometer data from the two cycles were available from 4 200 participants (rounded to the nearest 10 as per Statistics Canada confidentiality requirements) and were included in the current study. The mean scores (1 to 5, from better to worst) of participants on self-perceived mental health, self-perceived general stress and self-perceived work-related stress were respectively 1.97 (Standard error [SE] = 0.32, Mdn = 2.01), 2.78 (SE = 0.49, Mdn = 2.70) and 2.98 (SE = 0.49, Mdn = 2.90). Descriptive analysis also revealed a normal distribution both for general and work-related stress but a right-skewed one for the mental health scale. We observed that 31% report a high level of perceived work-related stress (4 = 25%; 5 = 6%) versus 20% (4 = 17%; 5 = 3%) and 5% (4 =1%; 5 = 1%) concerning perceived general stress and mental health, respectively. The mean age was 44 years (SE = 0.13) and 50.3% were women. Two thirds of the sample was in a relationship. On average, participants spent 21.3 (SE = 0.4, Mdn = 15.4) minutes per day of MVPA. Data from 14 univariate outliers were excluded because of extreme accelerometer data. Weighted characteristics of the study population are summarized in Table 1.

**Table 1.** Weighted characteristics of the *study population* (in millions) from cycles 1 and 2 of the Canadian Health Measures Survey (CHMS)

1 and 2 of the Canadian Health Measures Survey (CHMS)
|  | **N (%) or Mean ± SE** |
| --- | --- |
| Age | 44.96 ± 0.13 |
| Sex |  |
| Male | 1 2303 047 (49.82) |
| Female | 1 2391 325 (50.18) |
| Cotinine level (ng/mL) | 246.95 ± 18.054 |
| Education % |  |
| Less than high school diploma | 2 465 066 (9.98) |
| High school or work school | 7 924 541 (32.09) |
| College or university | 7 209 172 (29.19) |
| Not stated | 7 095 594 (28.73) |
| Household incomes (Canadian dollars) |  |
| <20k | 1 665 713 (6.75) |
| 20-<40k | 4 320 186 (17.49) |
| 40-<60k | 4 264 498 (17.27) |
| 60-<80k | 4 048 150 (16.39) |
| 80-<100k | 3 231 493 (13.09) |
| ≥100k | 7 164 333 (29.01) |
| Marital status |  |
| Alone | 8 570 418 (34.71) |
| In a relationship | 16 123 955 (65.29) |
| Body mass index (kg/m<sup>2</sup>) | 26.39 ± 0.19 |
| Leisure-time physical activity LTPA (METs-hour/day) | 1.84 ± 0.0562 |
| Moderate-to-vigorous physical activity MVPA (min/day) | 20.46 ± 0.76 |
| Total volume of PA (counts/day) | 190.47 ± 2.86 |
| Wear time of accelerometer (hours/day) | 13.73 ± 0.05 |
| Seasons of assessment |  |
| Autumn | 8 414 385 (34,07) |
| Spring | 5 491 546 (22,24) |
| Summer | 4 994 525 (20,23) |
| Winter | 5 793 916 (23,46) |
| Self-perceived mental health | 1.97 ± 0.32 |
| Excellent | 8 462 723 (34.52) |
| Very good | 9 682 090 (39.49) |
| Good | 5 396 853 (22.01) |
| Fair | 973 154 (3.97) |
| Poor | 179 553 (0.73) |
| Self-perceived general stress | 2.78 ± 0.49 |
| Not at all stressful | 2 363 075 (9.57) |
| Not very stressful | 6 545 954 (26.51) |
| A bit stressful | 10 764 333 (43.59) |
| Quite a bit stressful | 4 320 187 (17.49) |
| Extremely stressful | 700 824 (2.84) |
| Self-perceived work-related stress (WS) | 2,98 ± 0.49 |
| Not at all stressful | 2 212 523 (8.96) |
| Not very stressful | 4 855 196 (19.66) |
| A bit stressful | 10 153 007 (41.11) |
| Quite a bit stressful | 6 064 663 (24.56) |
| Extremely stressful | 1 408 983 (5.71) |
| Self-perceived health |  |
| Excellent, very good or good | 22 088 798 (89.45) |
| Fair or poor | 2 605 574 (10.55) |

### Shapes of association between exposures and outcomes

Figure 1 presents GAM results from adjusted models of WS as a respective function of MVPA and LTPA. The lines show the smoothed function from GAM for the two predictors and the dotted lines indicates the 95% confidence interval. Models including LTPA as the main predictor are also adjusted for MVPA since the Pearson correlation between the two measures is only of .32. Both LTPA and MVPA were significantly associated with self-perceived WS. A visual examination of the plots indicated nonlinear associations between both PA modalities and self-perceived WS.

*MVPA and WS*. A significant curvilinear relationship between MVPA and WS (p < 0.001, Adjusted R2 = 0.04) with an optimal duration of 90 minutes/day (1 hour 30 minutes) was found, decreasing WS by at least 0.5 and at best 1.0 scale point (respectively top and bottom of the confidence interval) compared with an absence of MVPA.

*LTPA and WS*. Concerning LTPA, a significant association with WS (p < 0.001, Adjusted R2 = 0.04) was observed with a more complex shape. The graph shows a bumpy fluctuation pattern until a drop at 8.5 MET/day, which means that an hour per day spent doing a leisure-time physical activity of 8.5 METs-hour is associated with the lowest WS level.

**Figure 1:** Dose-response relationships between WS-MVPA and WS-LTPA. Legend: WS = Work-related stress (Not at all stressful ‘ 1’, Not very stressful ‘2’, A bit stressful ‘3’, Quite a bit stressful ‘4’, Extremely stressful ‘5’), MVPA = Moderate and Vigorous Physical Activity, LTPA = Leisure-time physical activity. Lines represent smoothed means using generalized additive model, with short dashed lines representing 95% CIs and long dashed lines the respective optimum levels. Each model is weighted and adjusted for age, sex, daily smoking, household income, marital status, education levels, accelerometer wear time, season, income, body mass index, self-perceived general stress and self-perceived mental health.

## DISCUSSION

### Key findings

Based on a nationally representative sample of workers (n = 4 200), the aim of this study was to characterize the dose-response relationship between WS and two categories of physical activity, namely LTPA and MVPA.

Almost one third of our sample reported a high level of WS. Interestingly, there are many more people who are highly stressed by their work (WS, 31% of the sample) than by life in general (general stress, 20%), underscoring the importance of addressing this source of stress in particular. With respect to the dose-response analysis, significant results show that the daily optimal levels for reducing WS seem to be around 90 minutes for MVPA and 8.5 METs-hour in daily energy expenditure for LTPA, even after controlling for sociodemographic and psychosocial variables that are known to be related to WS (e.g. general stress). As a guide, MVPA ranges from moderate-intensity activities (3-6 METs), such as brisk walking, to vigorous-intensity activities (> 6 METs), such as carrying heavy loads or shovelling sand. LTPA worth 8.5 METs include, for example, backpacking, bicycling (25 km/h), handball, jogging (9 km/h) or swimming (3 km/h).^28,29^

Should be remembered that optimal doses of MVPA and LTPA do not necessarily represent actual daily values, but rather a mean (calculated over a one-week accelerometer monitoring period for MVPA and a three-month recall period for LTPA). It follows that different distributions of PA frequencies (e.g. more PA during the weekend and less during the weekday) could be just as beneficial in reducing WS. Interestingly, lower PA doses (see figure 1) may also provide benefit albeit to a lesser extent. Indeed, compared to a physically inactive person, engaging in 30 minutes of MVPA a day is associated with reduction in WS, which is slightly more than the World Health Organization recommendations of 150 minutes of MVPA throughout the week (≈ 20 minutes a day).^30^

Regarding LTPA, although its bumpy dose-response relationship with WS is relatively difficult to interpret, first signs of reduction in WS seem to appear from the moment the METs-hour exceeds zero, stressing the relevance of merely practising a LTPA, regardless of the energy expenditure. This is in line with previous studies which have shown that people who engaged in more PA reported lower WS and fewer burnout symptoms,^12,18^ yet without having established a dose-response relationship.

Taken together, these findings align with the tenet that getting some PA is better than none. In this respect, a growing body of literature indicates that even small increases of daily MVPA well below the optimum are nevertheless positively associated with mental health,^31,32^ although they did not measure WS.

### Strengths and limitations

To the best of our knowledge, this is the first Canadian nationally representative study to investigate the specific dose-response relationship between WS and two relevant categories of PA, namely MVPA and LTPA (N = 4 200). This study has the merit of shedding light on the optimal levels of MVPA/LTPA associated with a lower level of stress. Furthermore, those results are based on solid measure as MVPA was assessed using an Actical accelerometer.

The above findings notwithstanding, there are some limitations to this study that are worth noting. First, as it is often the case with large representative national surveys including several hundred questions, WS has been measured with a single-item scale that could be interpreted as equivocal. Thus, even though WS is nowadays negatively connoted and most often attributed to problematic psychosocial working conditions such as high job demands,^33^ it cannot be ruled out that WS could also have been interpreted by some respondents as “good stress” (eustress), which could explain the low effect size in the dose-response analysis.^34^ In fact, a stressor can also be perceived as a positive challenge if the individual thinks he has the abilities to overcome the stress-induced demands.^35^ In this case, the stressor may become a source of positive feelings^36^ and the neurophysiological response associated with eustress will be different from that triggered by distress.^37^ In light of this consideration, we strongly advocate that a single-item measure of WS would gain definitional clarity by simply giving the participant a unambiguous definition of “bad” work-related stress (work-related distress) in the instructions, such as “a particular relationship between the person and the environment that is appraised by the person as taxing or exceeding his or her resources and endangering his or her well-being”.^2^

The other limitation lies in the cross-sectional design preventing from establishing the direction of causality for the association between PA and WS. However, this concern is mitigated by prospective cohort studies indicating a protective effect of PA against the emergence of mental disorders.^38^

### Conclusion

Summing up the results, there are three things to keep in mind to help people clear their mind of WS: (1) one hour a day on average spent doing a LTPA of 8.5 METs-hour is associated with the highest benefits, (2) the optimal average daily dose of MVPA is around 90 minutes, and (3) first signs of reduction in WS appear long before optimum are reached (e.g. as soon as 30 minutes of MVPA), stressing the relevance of merely practising a LTPA/MVPA regardless of the dose. Drawn from a nationally representative sample, these findings are an important practical contribution, yet the pathways by which PA intervene in reducing WS are far from being fully understood. In this vein, further research may want to investigate the specific mechanisms underlying the relationship between WS and MVPA/LTPA. Ultimately, in applied sciences, moving from a merely probabilistic relationship to a mechanistic explanation is essential to more targeted and effective interventions.^39,40^

## Data Availability

Data are available upon request at Quebec inter-University Centre for Social Statistics (QICSS).

## Notes

### Competing Interest Statement

The authors have declared no competing interest.

### Funding Statement

Data access and analysis were sponsored by the Quebec inter-University Centre for Social Statistics (QICSS).

